# COVID-19 cases and hospitalizations averted by case investigation and contact tracing in the United States

**DOI:** 10.1101/2021.11.19.21266580

**Authors:** Gabriel Rainisch, Seonghye Jeon, Danielle Pappas, Kimberly D. Spencer, Leah S. Fischer, Bishwa B. Adhikari, Melanie M. Taylor, Bradford Greening, Patrick K. Moonan, John E. Oeltmann, Emily B. Kahn, Michael L. Washington, Martin I. Meltzer

**Affiliations:** CDC COVID-19 Response Team; CDC’s Epidemiology and Laboratory Capacity (ELC) Program

**Keywords:** COVID-19, case investigation, contact tracing, transmission, cases averted, hospitalizations averted, modeling

## Abstract

**Importance:** Evidence of the impact of COVID-19 Case Investigation and Contact Tracing (CICT) programs is lacking. Policymakers need this evidence to assess its value.

**Objective:** Estimate COVID-19 cases and hospitalizations averted nationwide by US states’ CICT programs.

**Design:** We combined data from US CICT programs (*e*.*g*., proportion of cases interviewed, contacts notified or monitored, and days to case and contact notification) with incidence data to model CICT impacts over 60 days period (November 25, 2020 to January 23, 2021) during the height of the pandemic. We estimated a range of impacts by varying assumed compliance with isolation and quarantine recommendations.

**Setting:** US States and Territories

**Participants:** Fifty-nine state and territorial health departments that received federal funding supporting COVID-19 pandemic response activities were eligible for inclusion. Of these, 22 states and 1 territory reported all measures necessary for the analysis. These 23 jurisdictions covered 42.5% of the US population (140 million persons), spanned all 4 census regions, and reported data that reflected all 59 federally funded CICT programs.

**Intervention:** Public health case investigation and contact tracing

**Main Outcomes and Measures:** Cases and hospitalizations averted; percent of cases averted among cases not prevented by vaccination and other non-pharmaceutical interventions (other NPIs).

**Results:** We estimated 1.11 million cases and 27,231 hospitalizations were averted by CICT programs under a scenario where 80% of interviewed cases and monitored contacts, and 30% of notified contacts fully complied with isolation and quarantine guidance, eliminating their contributions to future transmission. As many as 1.36 million cases and 33,527 hospitalizations could have been prevented if all interviewed cases and monitored contacts had entered into and fully complied with isolation and quarantine guidelines upon being interviewed or notified. Across all scenarios and jurisdictions, CICT averted a median of 21.2% (range: 1.3% – 65.8%) of the cases not prevented by vaccination and other NPIs.

**Conclusions and Relevance:** CICT programs likely had a substantial role in curtailing the pandemic in most jurisdictions during the winter 2020-2021 peak. Differences in impact across jurisdictions indicate an opportunity to further improve CICT effectiveness. These estimates demonstrate the potential benefits from sustaining and improving these programs.

**KEY POINTS:** *Question:* What were the health impacts of COVID-19 case investigation and contact tracing programs (CICT) in the US?

*Findings:* By combining CICT program data from 22 states and 1 territory with mathematical modeling, we estimate CICT averted between 1.11 to 1.36 million cases and 27,231 to 33,527 hospitalizations over 60 days during the height of the pandemic (winter 2020-21). The upper estimate assumes all interviewed cases and monitored contacts complied with isolation and quarantine guidelines, while the lower estimate assumes fractions of interviewed cases and monitored or notified contacts did so.

*Meaning:* CICT programs likely played a critical role in curtailing the pandemic.

## INTRODUCTION

Reducing exposure to persons with communicable diseases through isolation and quarantine are basic tenets of transmission prevention. Public health programs regularly conduct case investigation and contact tracing (CICT) as a means of notifying persons infected with or exposed to communicable diseases and, often, of their need to isolate or quarantine. However, evidence of CICT’s role in mitigating the COVID-19 pandemic thus far is lacking.^1^ We recently showed, using data from 14 US jurisdictions (five states and nine local health districts), that CICT programs were effective at reducing SARS-CoV-2 transmission.^2^ Despite these findings, the impact and consequent value of CICT remains controversial.^3-6^ Some claim that the benefits are limited due to difficulty in scaling up services during COVID-19 case surges, or community reticence to participate in CICT, curtailing meaningful engagement between health departments and cases and their close contacts.^3, 6, 7^ Between June 2020 and March 2021, the US Centers for Disease Control and Prevention (CDC) distributed more than $40 billion to state, local, and territorial health departments to support COVID-19 response activities, with a notable portion directed toward CICT activities.^5^ A national review of these efforts from November 2020 to March 2021, indicates that upwards of 42,000-55,000 case investigators and contact tracers (per month) interviewed 9.1 million cases and identified and sought to notify 10.7 million contacts.^5^ Given the unprecedented funding and effort surrounding CICT and continuing debate surrounding its value, we sought to present an expanded profile of national CICT impacts at its busiest point of the pandemic. This study uses data from 22 states and 1 US territory, with a combined population of 140 million people, during the peak months of COVID-19 incidence in the US - November 2020 through January 2021.

## METHODS

We used CDC’s COVIDTracer Advanced modeling tool ^8^ in combination with data from CICT programs to estimate cases and hospitalizations averted by CICT activities among states and territories funded by CDC’s Epidemiology and Laboratory Capacity for Prevention and Control of Emerging Infectious Diseases (ELC) program. We focused on the 60-day period from November 25, 2020 to January 23, 2021.

### Data

Sixty-four health departments receiving CICT funding report to CDC’s ELC program monthly on the performance of their CICT programs.^5^ We used reported metrics from each jurisdiction to derive its CICT effectiveness for the 60-day analysis period: the proportion of cases and contacts that entered into isolation and quarantine because of CICT efforts, and the days required to do so (Supplement 1, *Case Investigation and Contact Tracing Effectiveness* section). Reported metrics used to calculated CICT effectiveness include the proportions of cases interviewed, contacts notified or monitored, and number of days from testing to case and contact notification. A summary of these data and assumptions used to calculate a range of CICT effectiveness values for each jurisdiction are detailed further below and in Supplement 1 (*Case Investigation and Contact Tracing Effectiveness* section and eTable 4). We limited our analysis to those jurisdictions that reported all the required metrics and passed our data quality checks (*e*.*g*., the number of contacts identified ≥ number cases that provided at least one contact, the number of contacts identified ≥ contacts notified; eFigure 3 in Supplement 1).

### Model Use

COVIDTracer Advanced is a spreadsheet-based epidemiological model that illustrates the spread of COVID-19 and impact of interventions in a user-defined population. The tool allows users to attribute reductions in transmission to either CICT or to a combination of vaccination and all other non-pharmaceutical interventions (other NPIs), such as facemask policies, large gathering restrictions, and school/business closures (Supplement I, *COVIDTracer Advanced Model* section). Estimates of reductions in transmission from CICT were obtained by first entering each jurisdiction’s CICT effectiveness into COVIDTracer Advanced. After inputting the CICT effectiveness values for a jurisdiction, we estimated reductions in transmission due to other NPIs and any inceptive vaccination efforts. We accomplished this by “fitting” the curve of cumulative cases modeled by COVIDTracer Advanced to the jurisdiction’s reported cases ^9^ by altering the percentage reduction in transmission ascribed to vaccine and other NPIs. The value that minimized the deviation (mean-squared error) between the fitted and reported case curves was our estimated effectiveness of vaccine and other NPIs (eTable 5 in Supplement 1). Then we “switched off” CICT (by setting CICT effectiveness to zero) while maintaining the estimated effectiveness of vaccine and other NPIs. This simulated what would have happened if CICT had not occurred. Readers can replicate this process with the provided Special Edition version of COVIDTracer Advanced (Supplement 2) and accompanying instructions (Supplement 1).

### Outcome Measures

Estimates of CICT-averted cases were obtained by taking the difference between the model-simulated curve without CICT and jurisdictions’ actual cumulative cases. We also calculated averted hospitalizations by multiplying the estimated number of averted cases by age-stratified infection-to-hospitalization rates (eTable 3 in Supplement 1). In addition to calculating the absolute number of cases and hospitalizations averted by CICT in each jurisdiction, we calculated two measures of CICT impact to allow comparison among jurisdictions: 1) averted cases and hospitalizations per 100,000 population, and 2) the proportion of cases or hospitalizations averted by CICT out of the remaining cases, after accounting for the impact of vaccination and other NPIs. This latter measure may be interpreted as the number of cases (or hospitalizations) averted by CICT among every 100 cases (or hospitalizations) that were not prevented by vaccination and other NPIs. Finally, we grouped jurisdictions by their US Census Region and compared the group medians of cases averted per 100,000 population to assess whether CICT impact differed among regions.^10^

### Range of estimates

Jurisdictions did not report the proportions of cases that effectively isolated and contacts that correctly quarantined. Absent compliance data, we generated a range of averted cases and hospitalizations to circumscribe the possible impact of CICT. High estimates were calculated by assuming all of the cases a jurisidiciton interviewed and all of the contacts it actively monitored fully complied^2^ with CDC-recommended isolation and quarantine guidelines (Table 1).^11^ In our high estimate scenario, we also assumed that contacts who were notified but not actively monitored did not quarantine. That is, we assumed that the cases and contacts CICT programs engaged either fully complied or not at all in this scenario. To produce our low estimates, however, we altered the effect of CICT program’s engagement by lowering the proportions of cases/contacts entering isolation or quarantine based on values derived from the literature (*Isolation/Quarantine Compliance* section in Supplement 1).^7, 12, 13^ Specifically, we assumed 80% of cases that completed interviews, 80% of monitored contacts, and 30% of notified contacts (that were not actively monitored) fully complied with isolation and quarantine guidance (Table 1). In our model, and irrespective of the above scenarios, infected individuals may transmit to others until interactions with their health department prompt them to isolate or quarantine. Also, both estimates do not include unmeasured changes in behavior from sources of information other than CICT programs (*e*.*g*., cases directly informing their contacts) as measuring the influence of such factors are beyond our estimation goals.

**Table 1.** Assumed proportions of confirmed cases and their contacts that effectively isolated or quarantined in each analysis scenario.

|  | **Scenario 1**<br>**(low estimate)** | **Scenario 2**<br>**(high estimate)** | **Sensitivity Analysis**<br>**(maximal estimate)** |
| --- | --- | --- | --- |
| **Confirmed Cases that completed interviews** | 80% | 100% | 100% |
| **Confirmed Cases that did not complete interviews<sup>a,b</sup>** | 0% | 0% | 0% |
| **Contacts that were notified and monitored** | 80% | 100% | 100% |
| **Contacts that were notified but not monitored** | 30% | 0% | 100% |
| **Contacts that were notified by their health department<sup>b</sup>** | 0% | 0% | 0% |

**Note:** Each row is a mutually exclusive group of cases or contacts. The sum of each row (or column) does not add up to 100%, as the numbers represent the assumed compliance within each group. 0% compliance means none of the cases or contacts in a group isolated or quarantined effectively. 100% means all of the cases or contacts in a group isolated or quarantined effectively after being interviewed or notified.
<sup>a</sup> Includes cases that weren't reached and those that were reached but who did not agree to be interviewed.
<sup>b</sup> Compliance was set to zero for these case/contact group categories because any transmission reductions from quarantine and isolation are not attributable to direct interactions with health department's CICT staff, and, therefore outside of the scope of this analysis. Their inclusion here is to help distinguish between the various cases/contacts types.

We also conducted a sensitivity analysis to evaluate the potential that compliance with quarantine and isolation guidance exceeded those assumed here. Absent data indicating quarantine compliance among all contacts, we chose to evaluate the impact of CICT assuming maximum community cooperation. Therefore, we assumed a scenario in which all interviewed cases and all contacts notified of their exposure fully complied with CDC-recommended quarantine guidelines.

## RESULTS

Twenty-two US states and one territory met our data requirements for inclusion in the analysis (Figure 3 in Supplement 1). These 23 jurisdictions had a combined population of approximately 140 million persons, covering 42.5% of the entire US population and all 4 census regions.^10^ There were 5,269,390 total cases reported across these jurisdictions during our 60-day study period. Jurisdictions in our analysis reported metrics (% of cases interviewed and contacts notified, and contact notification speed) that were similar to those reported by all 64 federally funded CICT programs (eTable 4 in Supplement 1).

We estimated that CICT averted 1.11 to 1.36 million cases and 27,231 to 33,527 hospitalizations from November 25, 2020 to January 23, 2021 across all 23 jurisdictions analyzed (Figure 1 and eFigure 4 and eTable 5 in Supplement 1). The lower estimates assume fractions of interviewed cases and contacts complied with isolation and quarantine guidelines, while the upper estimates assume all interviewed cases and monitored contacts did so (Table 1). The median number of estimated cases averted per 100,000 population ranged from 704 in the scenario with lower overall compliance, to 895 in the scenario with higher overall compliance. After accounting for the impact of vaccination and other NPIs, the median estimate of the percent of cases averted was 19.1% (range: 1.3 – 65.8%) in the low compliance scenario and 23.5% (range: 1.6 – 58.7%) in the high compliance scenario (eTable 5 in Supplement 1).

**Figure 1.**
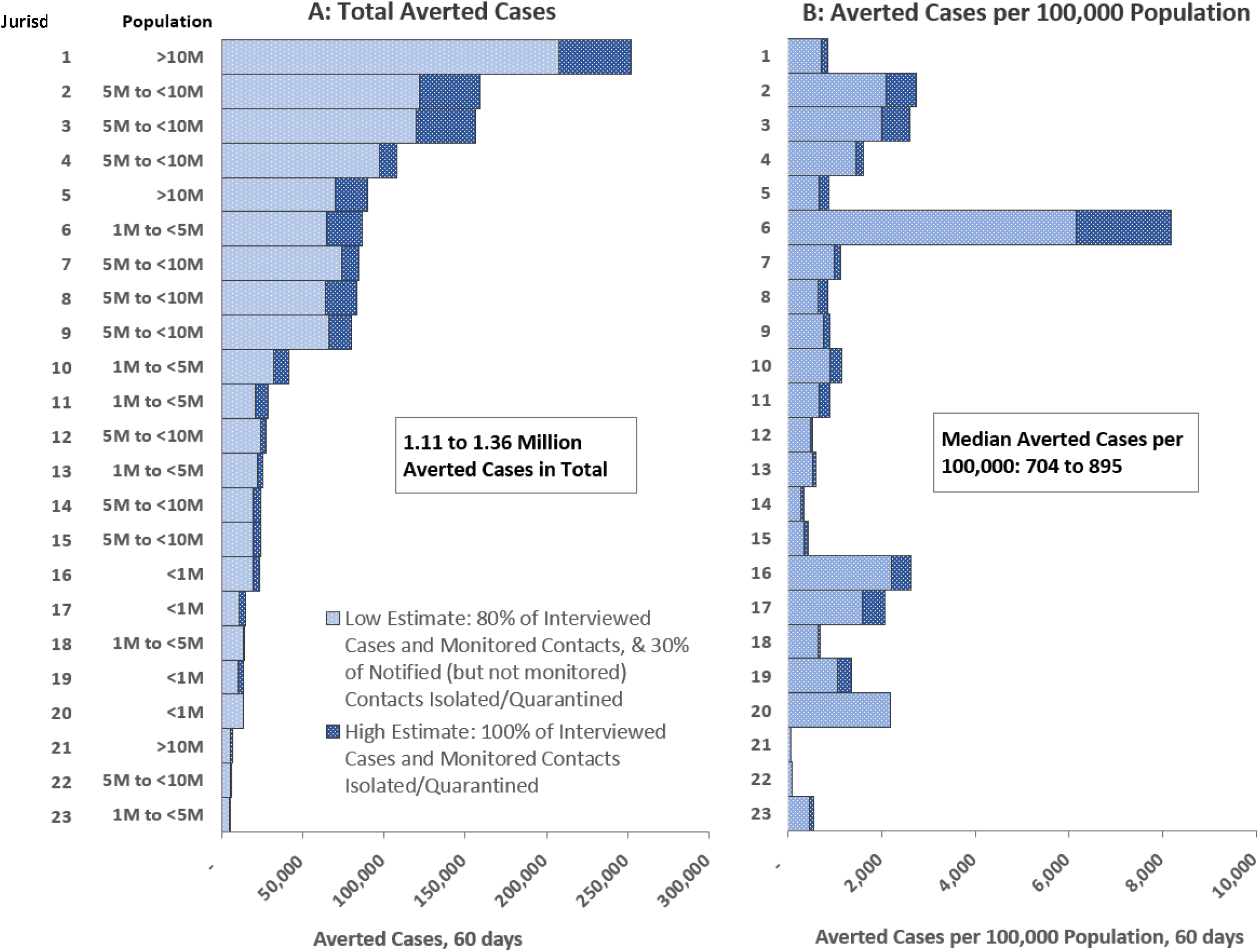
Estimated COVID-19 Cases Averted by Case Investigation and Contact Tracing, by Jurisdiction, November 25, 2020-January 23, 2021 (60 days)

On average, the number of estimated cases averted was greater among jurisdictions with larger populations, with more jurisdictions in the top half of Figure 1, Panel A having greater populations than those in the bottom half: the median population size of jurisdictions in the top half was 6.4 million (IQR: 4.8 – 9.2 million) and 3.2 million (IQR: 1.0 – 6.0 million) for the bottom half. However, per our estimates, CICT programs in jurisdictions with smaller populations often averted more cases on a per population basis (Figure 1, Panel B). Jurisdictions in the smallest population category (less than 1 million) averted the most cases per population (median: 1,714 – 1,875 per 100k population); more than twice the overall median (704 - 895 per 100k population).

We estimate that jurisdictions in the Midwest US averted the most cases on a per population basis because of CICT, averting between 1,444 cases per 100,000 population (in our low compliance scenario) and 1,600 (in our high compliance scenario) (Table 2). CICT programs among jurisdictions in the Western US were the least effective by our estimates, averting 488 cases per 100,000 (in our low compliance scenario) to 568 (in our high compliance scenario). When we maximized compliance among interviewed cases and notified contacts (Table 1), we estimated that CICT averted 1.72 million cases and 42,2631 hospitalizations (approximately 26% greater than our high baseline estimate) across the 23 jurisdictions during the 60-day study period.

**Table 2.**
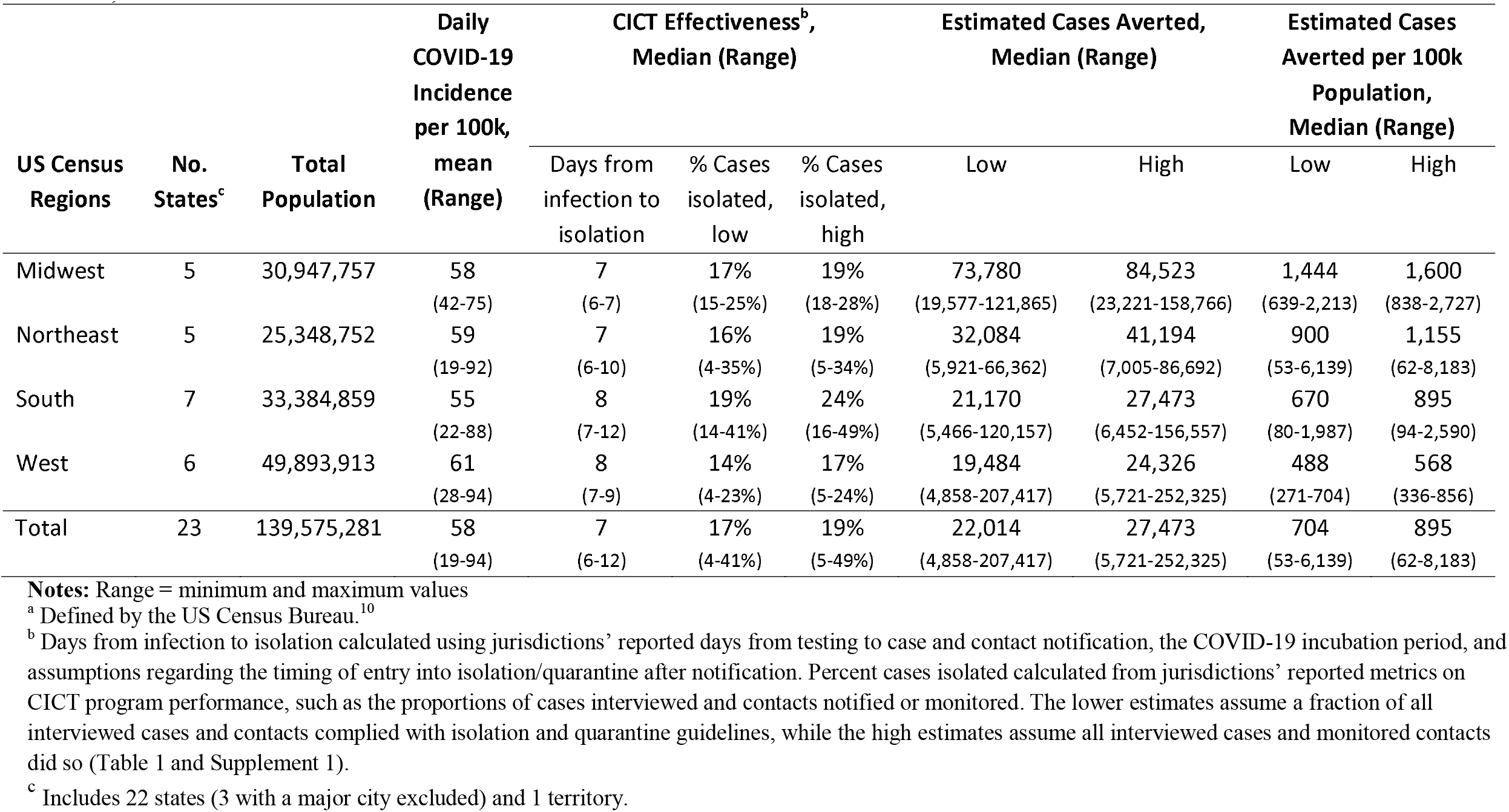
Case Investigation and Contact Tracing Effectiveness and Health Impacts, by US Census Region^a^ (from 11/25/20 – 1/23/21)

## DISCUSSION

We estimate that these 23 CICT programs potentially averted between 1.11 and 1.36 million cases and from 27,231 to 33,527 hospitalizations in a 60-day period during the height of the pandemic (winter of 2020-21). There were 5,269,390 total cases reported across these jurisdictions during the same 60-day period, suggesting that CICT may have reduced the COVID-19 burden by 17 to 21%. Our range of estimates reflect uncertainties regarding the proportions of cases and contacts that effectively isolated or quarantined as a result of interactions with their health departments. Despite this uncertainty, our estimates of CICT impact were substantial, with averted cases exceeding 1 million across the 23 jurisdictions in our low compliance scenario. Even our high estimates may underestimate the potential impacts in some jurisdictions because we assumed notified contacts did not quarantine unless actively monitored. Had all interviewed cases and notified contacts isolated and quarantined, as examined in our sensitivity analysis, these 23 CICT programs would have averted as many as 1.72 million cases and 42,2631 hospitalizations. While aspirational, these values quantify the potential benefit from maximimum public compliance with isolation and quarantine guidance. Finally, our estimates do not account for the indirect effects of CICT programs on transmission reductions. Due to their interactions with health department staff, cases and contacts may have additionally notified and motivated isolation/quarantining among close contacts whom, themselves, were not contacted by the CICT program.

While our aggregate estimated impact was sizeable, it was uneven across the jurisdictions: In the lowest performing jurisdiction, CICT averted just 1 out of every 100 remaining cases not prevented by nascent vaccination efforts and other NPIs, and as many as 66 cases in the highest performing jurisdiction. We also found that population size was correlated with our estimates of CICT impact. On average, our estimates suggest that jurisdictions with larger populations averted more cases, although this was expected given larger populations eligible for protection. Conversely, the smallest jurisdictions averted the most cases on a per-population basis. These results may reflect, in part, that smaller jurisdictions were able to rely on existing CICT staff who had community knowledge and experience connecting with the population, while the caseloads in larger jurisdictions required hiring temporary, less experienced staff. Also, on average, larger jurisdictions had a higher mean daily incidence per 100k population over the 60-day study period (albeit this correlation was weak: Pearson correlation coefficient = 0.25). A multivariable analysis, using data from several months of the pandemic, is needed to tease apart the effects of such factors. For example, population size alone cannot explain the variability in our estimated impacts. Jurisdictions 5, 6, and 7 were in different population categories (with jurisdiction 5 being at least 10 times larger than jurisdiction 6), but all three jurisdictions averted approximately 87,000 cases under our high compliance scenario (Figure 1, Panel A). Jurisdiction 6’s CICT program is also notable for averting the most cases per 100,000. This result reflects the jurisdiction’s success at interviewing cases (79% interviewed and >50% named at least 1 contact) and being among the fastest to notify contacts (6 days after cases were likely infected). We also found regional differences in CICT impact. Based on the median averted case estimates per 100,000 population, Midwest jurisdictions’ CICT programs performed the best, while CICT programs in Western jurisdictions were least impactful. Future studies exploring the potential reasons for these differences may consider incidence, factors affecting public acceptance of CICT (*e*.*g*., sociodemographic makeup and cultural norms), and aspects of program implementation (*e*.*g*., staffing levels and efficiency).

Our sizeable estimates of averted cases are partially due to the success of the analyzed CICT programs at suppressing the exponential growth in cases associated with uncontrolled transmission (2.5 new infections per case without interventions and almost all the population susceptible to infection [eTable 2 in Supplement 1]), compounded over approximately 10 generations of infection during our 60-day observation period. For example, at jurisdiction 1, where our estimates of the absolute impact of CICT was greatest, CICT was responsible for just a 3.0 to 3.5% reduction in new infections per case (eTable 5 in Supplement 1). However, jurisdiction 1 also had a very large burden of infectious cases at the start of our 60-day period and was one of the largest jurisdictions. This example shows that even when the percentage reduction in transmission from CICT is in the low single digits, when applied to large populations, the influence over multiple generations of cases is meaningful.

These results of state-level CICT impact have similarities and differences with our previous, mostly local-level study.^2^ The median percent of cases averted by CICT (after accounting for the impact of vaccination and other NPIs) in the previous study was double our estimates here: 42.3% (range 4-97%) among the 14-sites ^2^, versus 21.2% (range 1% – 66%) in this study. While this may be driven by factors correlated with population size previously mentioned, caseload differences and subsequent burden on local health departments during the periods analyzed likely contributed as well: the average daily COVID-19 incidence in this analysis period was 10 times greater than the previous study.^9, 14^ Finally, differences between studies may simply reflect the different samples of sites analyzed. Nevertheless, the wide range of impacts in both studies shows that variability in CICT program performance exists at both the state and local level. Our study has several strengths. Foremost, the breadth of data on CICT implementation enabled us to generate a profile of CICT impact for nearly half of the US. Further, this work can be replicated for other jurisdictions and time periods. The tool that we used, COVIDTracer Advanced ^8^, is provided (Supplement 2) and designed for use by practicing public health officials. By anonymizing jurisdictions and assessing the same time frame, we were able to present and compare the range of impacts among 23 CICT programs spanning the country. Jurisdictions assessing the value of CICT can consider this multi-jurisdictional analysis and conduct site-specific analyses using these methods to estimate prevention impact, guide local public health programming, and reflect on resource utilization (*e*.*g*., hospital beds).

Our study also has limitations. Jurisdictions’ self-reported CICT performance measures were not intended for this analysis. Although we employed the previously described data quality checks (eFigure 3 in Supplement 1), the reported measures that we used were likely influenced by differences in jurisdictions’ surveillance systems, CICT platforms and protocols (*e*.*g*., how they enrolled and monitored contacts). The extent to which these differences affected our results is unclear. We also only assess the impact over two months (60 days) of the pandemic and in 23 US jurisdictions. Results may differ for other periods (*e*.*g*., during the surge of the Delta variant and wider use of vaccine) and jurisdictions. Because cases were spiking across the entire US during the period that we analyzed and the vaccine had not yet been widely administered, it is likely that our estimates provide an upper limit of cases averted by CICT during the pandemic as of this writing (August 31, 2021). Finally, because we used statewide data, our results dilute potentially meaningful differences in CICT performance within jurisdictions (*e*.*g*., rural versus urban counties).

Our analysis combined primary implementation data with mathematical modeling to estimate the health impact of COVID-19 case investigation and contact tracing across nearly half of US state and territory CICT programs. The volume of estimated cases and hospitalizations averted underscores the critical role CICT programs play in curtailing the pandemic, while differences among jurisdictions illustrate the opportunities to further improve effectiveness. Case investigation and contact tracing remain CDC-recommended practices for personally communicating individualized prevention activities against COVID-19.^15^ This work quantifies for public health decision makers the benefits from sustaining and improving these programs.

## Supporting information

Supplement 2_COVIDTracer-Advanced-SpecalEdition

## Data Availability

All data are available upon request.

## Acknowledgements

Alison Stargel and Alvin Shultz with CDC’s Epidemiology and Laboratory Capacity (ELC) Program for assistance with data use and interpretation; and all of CDC’s Prevention and Control of Emerging Infectious Diseases Cooperative Agreement recipients, including state, local and territorial health departments, for the use of their CICT process metrics.

## SUPPLEMENT 1 – TECHNICAL APPENDIX

### COVIDTracer Advanced Model

COVIDTracer Advanced^1^ is a spreadsheet-based compartmental Susceptible-Exposed-Infectious-Recovered (SEIR) epidemiological model, which illustrates the spread of a pathogen, resultant disease, and impact of interventions in a user-defined population. Readers can download the tool and enter input values of their choosing, exploring the impact of scenarios and assumptions beyond those covered in this manuscript. To model the clinical progression and transmission of disease using COVIDTracer Advanced, we used the following definitions and assumptions. A “case” was defined as a person who has been exposed, infected and subsequently becomes infectious, regardless of the presence of clinical symptoms. We assumed that for the first 3 days after infection, cases do not infect others. During days 4–5 post-infection, cases are pre-symptomatic but shed virus in amounts that may infect others.^2-5^ During days 6–14, the infected person can be symptomatic and shedding virus, albeit during days 11–14 the risk of onward transmission is relatively low but non-zero (the complete infectivity distribution is given in Table A1). We assumed that approximately 40% of cases are asymptomatic during days 6-14 yet have a risk of onward transmission equal to 75% of symptomatic cases (Table A2) without vaccine or other NPIs.^5^ The model assumes homogeneous mixing among individuals and does not account for any age- or location-based heterogeneities in transmission.

**eTable 1.**
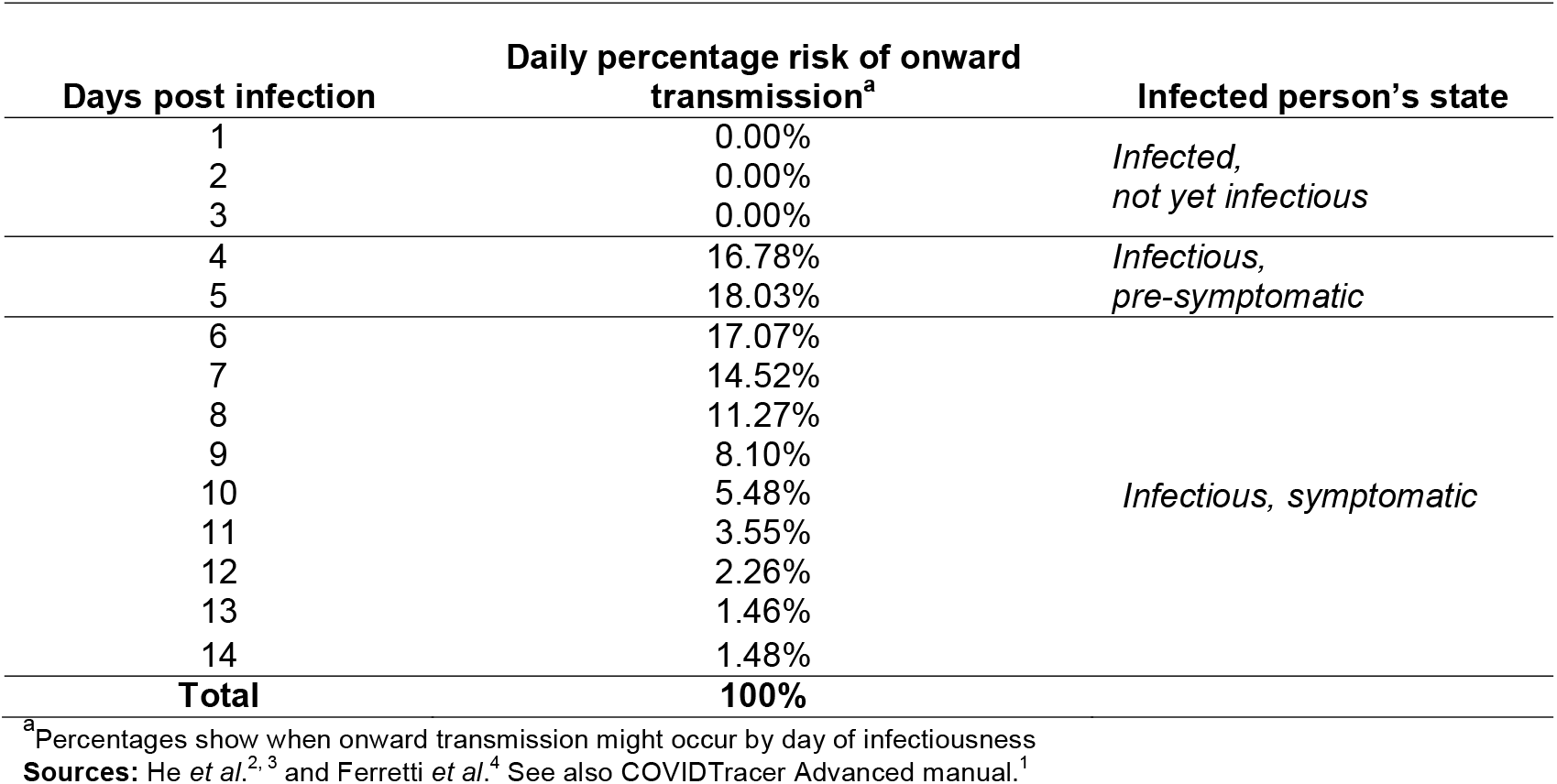
Daily percentage risk of transmission by infectiousness state and clinical symptoms.

**eTable 2.**
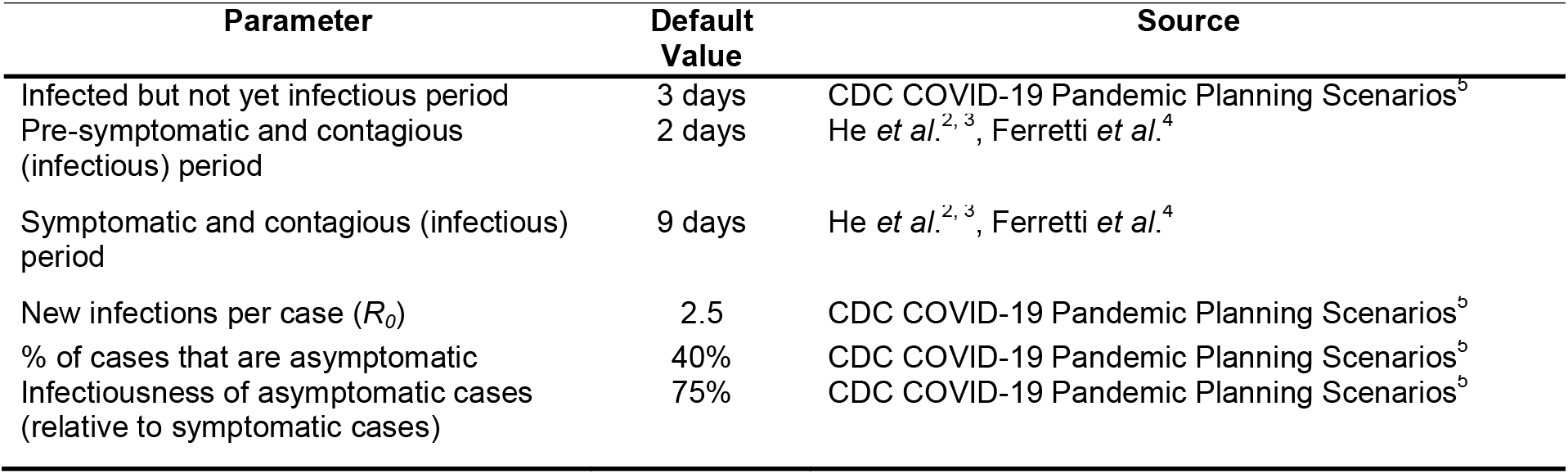
Epidemiological parameters, values, and sources.

**eTable 3.**
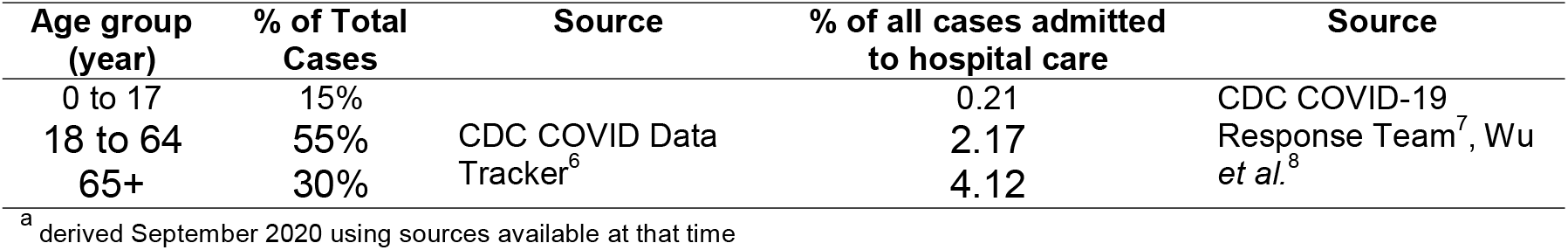
Assumed^a^ proportion of cases by age group and infection-to-hospitalization rate, default values in COVIDTracer Advanced and sources.

**eFigure 1.**
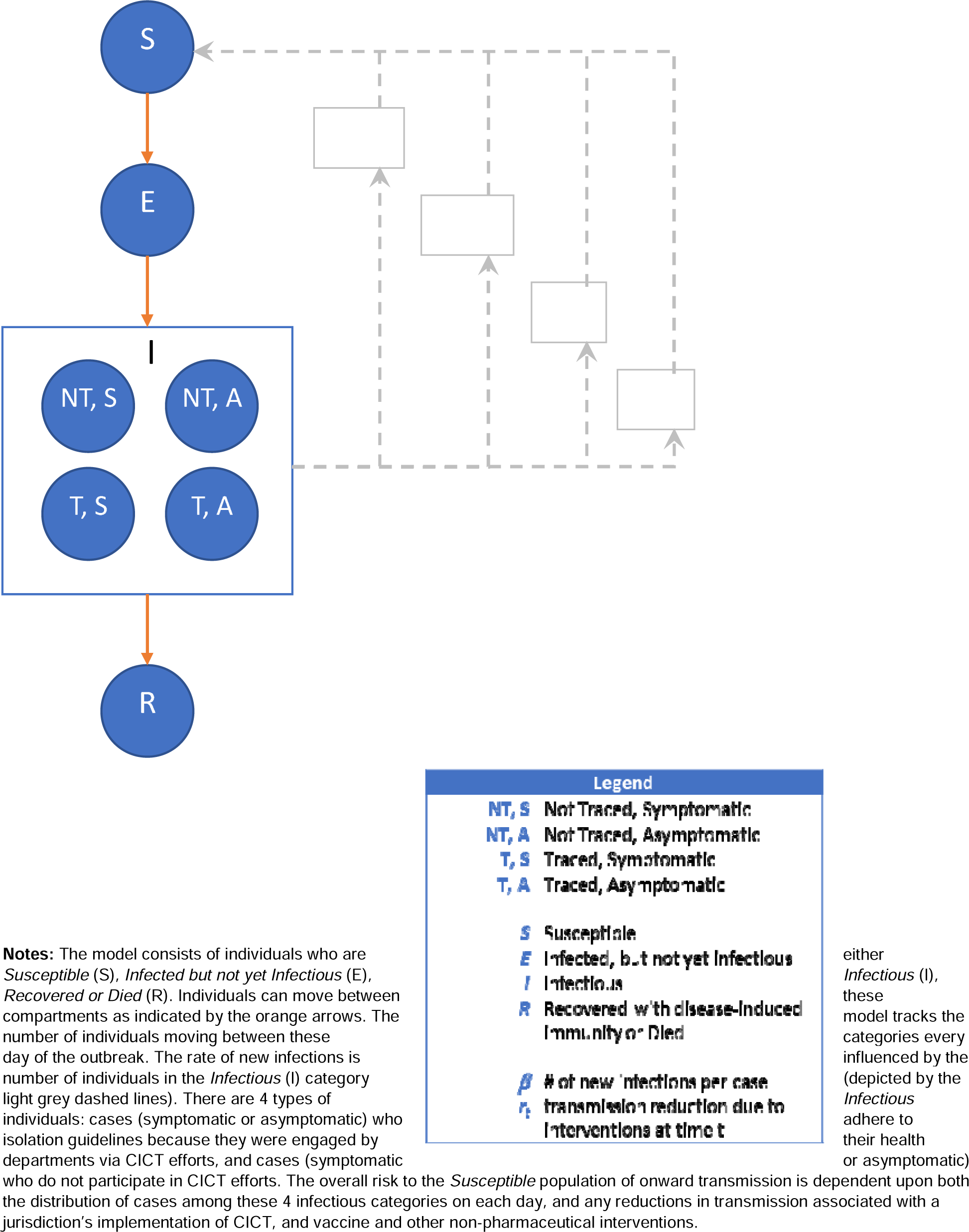
COVIDTracer Advanced Model Structure.

### Case Investigation and Contact Tracing Effectiveness

The effectiveness of case investigation and contact tracing is determined by the proportion of cases and their contacts that are effectively isolated and quarantined, preventing further transmission in the susceptible population. The duration of quarantine and isolation is described in CDC’s guidance.^9^ We assumed that confirmed cases are effectively isolated following case interviews. We further assumed that contacts are quarantined upon either contact notification or through active monitoring. If infected contacts are left undetected, they will infect additional contacts (on average, 2.5 new infections per infected contact during the period of the pandemic analyzed). Therefore, we calculated the average proportion of cases and contacts isolated and quarantined for each location as follows:

**Equation 1: Baseline Low Estimate**

80% of interviewed cases and monitored contacts, and 30% of notified contacts (who are not monitored), isolate or quarantine:

> Average proportion of cases and contacts (which become cases) isolated = [% Cases interviewed*0.8 + (*R*_*0*_ * % Contacts identified * (% Contacts monitored *0.8 + % Contacts notified but not monitored*0.3))] / (1+*R*_*0*_)

**Equation 2: Baseline High Estimate**

100% of interviewed cases and monitored contacts isolate or quarantine:

> Average proportion of cases and contacts (which become cases) isolated = [% Cases interviewed + (*R*_*0*_ * % Contacts identified * % Contacts monitored)] / (1+*R*_*0*_)

**Equation 3: Sensitivity Analysis (Maximum CICT Impact) Estimate**

100% of interviewed cases and 100% of contacts isolate or quarantine:

> Average proportion of cases and contacts (which become cases) isolated = [% Cases interviewed + (*R*_*0*_ * % Contacts identified * % Contacts notified)] / (1+*R*_*0*_)

where *R*_*0*_ is the assumed number of new infections per case without any interventions and when the population is entirely susceptible to infection (Table A2). The % Cases interviewed, % Contacts notified, and % Contacts monitored were metrics reported by jurisdictions and the % Contacts identified was an intermediate value calculated as the number of named contacts divided by the expected number of contacts per case:

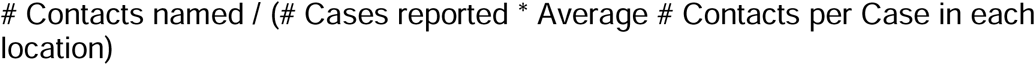

In addition, reducing the time from case identification to effective isolation is critical for case investigation and contact tracing to succeed. The longer the cases and contacts interact with the susceptible population, the greater the opportunity for onward transmission. In practice, cases with no known exposure are predominantly identified and isolated after symptom onset^3^, and cases with known exposures (*i*.*e*., contacts that eventually become infected cases) can begin quarantine upon contact notification (even potentially prior to symptom onset). We assumed asymptomatic cases can only be identified and isolated if they are notified through case investigation and contact tracing. For the purposes of our study, we assumed the proportions of cases with no known exposure and cases with known exposures were equal (*i*.*e*., 50/50 breakdown) because we did not have data on what prompted case identification in each location. Therefore, for each location the days to effective case isolation was determined by taking the average of the days to effective isolation between case groups with known and no known exposures. The time to effective case isolation for each of the two case groups was determined as follows:

For symptomatic cases with no known exposures (i.e., symptoms prompt identification): We assumed that cases experience a 5-day pre-symptomatic period (See Table A2), get tested the day after symptom onset (*i*.*e*., 6 days would had transpired since infection at the time of testing). We then obtained the number of days from testing to interview completion by taking the sum of jurisdictions’ reported “*Median days from specimen collection to case reporting to the health department (HD)”* and “*Median days from case report to the HD to case interview completion”*. We also assumed that confirmed cases begin isolation the day after their interview with the health department (*i*.*e*., we added 1 to the total obtained above). Our assumptions regarding the “next-day” timing of testing and entry into isolation are based on symptoms and notifications beginning or occurring throughout the day, with a sizeable portion occurring sufficiently late enough in the day to prevent testing and entry into isolation the same evening. This assumption takes into account practical considerations such as time needed to find a testing site and arrange an appointment, and for notified individuals to prepare to isolate (*e*.g., purchasing food or medications, setting up childcare, handling work or other commitments).

For cases with known exposures (i.e., those who were notified they were a contact and eventually became a case):

We first calculated the days from index case testing to their exposed contacts’ notification by summing jurisdictions’ reported “*Median days from specimen collection to case report to the HD”*, “*Median days from case report to the HD to the case interview completion”*, and “*Median days from case interview completion to contacts notification”*. We assumed that contacts begin quarantine the day after receiving exposure notification from their health department (*i*.*e*., we added 1 to the sum above). The “next-day” timing of entry into quarantine is based on the same practical reasoning as cases needing time to prepare to isolate once notified (described above). We then used the resultant sum from the procedure above to estimate the time (in days) from exposure to quarantine for contacts. Because we did not have information on when exposures actually occurred for contacts, we assumed that these individuals’ exposures occurred at the midpoint of their potential exposure window (in days). We identified the earliest date in this window as the first day of infectiousness among cases to which contacts were exposed. Based on our assumed 5-day pre-symptomatic period for symptomatic cases (described above), this was two days prior to the symptom onset date in cases exposing the contact. We identified the latest possible exposure as the date the cases exposing them were interviewed by the health department (because they began isolation the next day). See both “Contacts” rows in Figure A2 for a visual depiction of this timeline.

**eFigure 2.**
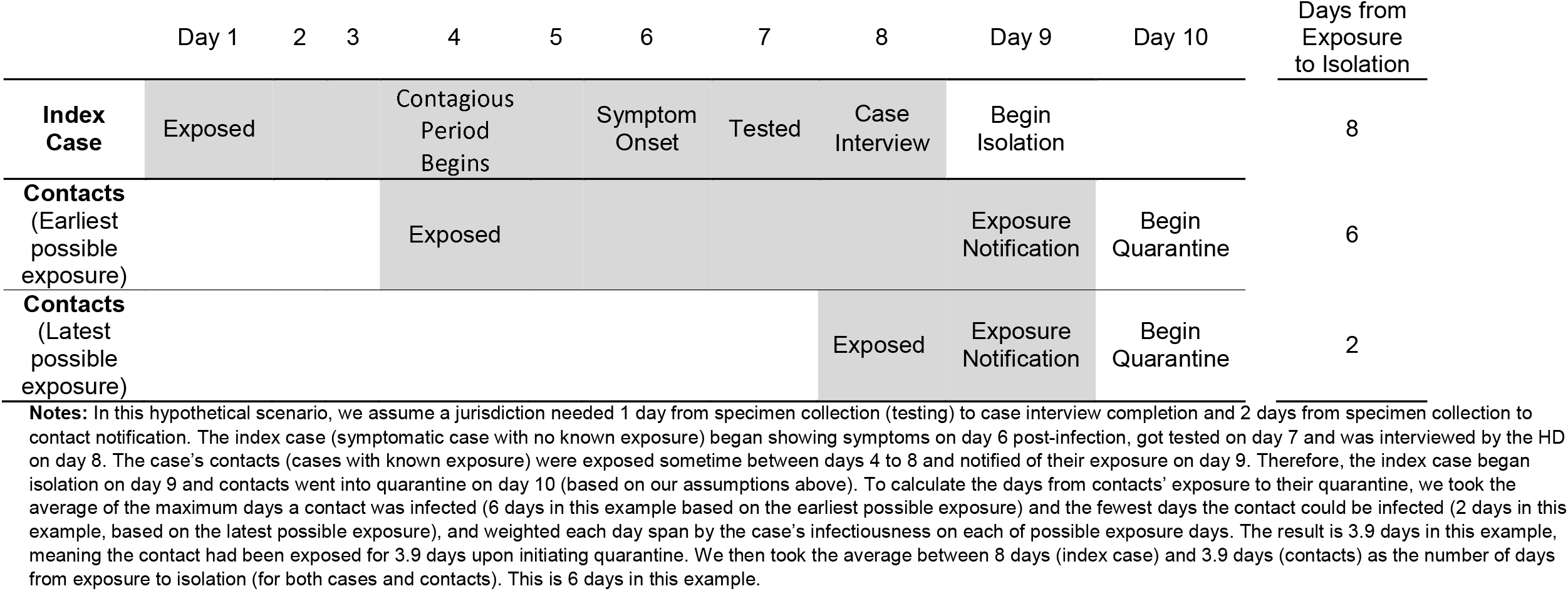
Illustrative example of the timing of COVID-19 case isolation and quarantine of contacts.

The days between cases with known exposures becoming infected and their exposure notification can vary from what we assumed. For example, cases may take longer to become symptomatic, or get tested the same day that they become symptomatic or begin their isolation on the same day as their results notification. Similarly, contacts who become cases may be exposed earlier or later than we assumed and may make up a larger or smaller share of the case pool. Readers interested in more detail of the influence of varying our assumed time to case isolation may wish to see Table A6 in the Technical Supplement of our 14-site study on CICT impact, containing results of a sensitivity analysis examining this topic in those jurisdictions.^10^

ELC-funded jurisdictions also reported the *Number of contacts that were notified within 1 day of case interview*, the *Number of contacts that were notified between 1-3 days after case interview*, and the *Number of contacts that were notified within 3 or more days after case interview*. We used these additional data elements as a quality check (Figure A3) of the reliability of jurisdictions’ reported median values regarding notification timing (described above). We did this by calculating the lower limit of the average number of days from case interview to contact notification as follows:

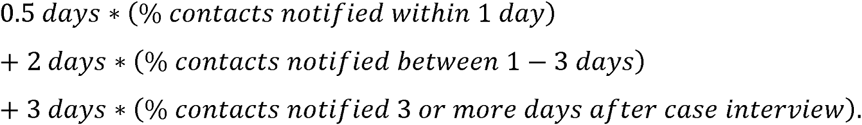

This metric assumes that all contacts were notified within 3 days of the case interview. We used this metric to exclude jurisdictions from the analysis (*i*.*e*., deemed reported data unreliable) when the lower limit of the average time to contact notification was greater than our calculated time to contact notification using reported median days AND the proportion of contacts that were notified 3 or more days after case interview was less than 10% of total contacts (*i*.*e*., too few to exert enough influence on the average lower limit for it to plausibly exceed the median-based value).

**eFigure 3.**
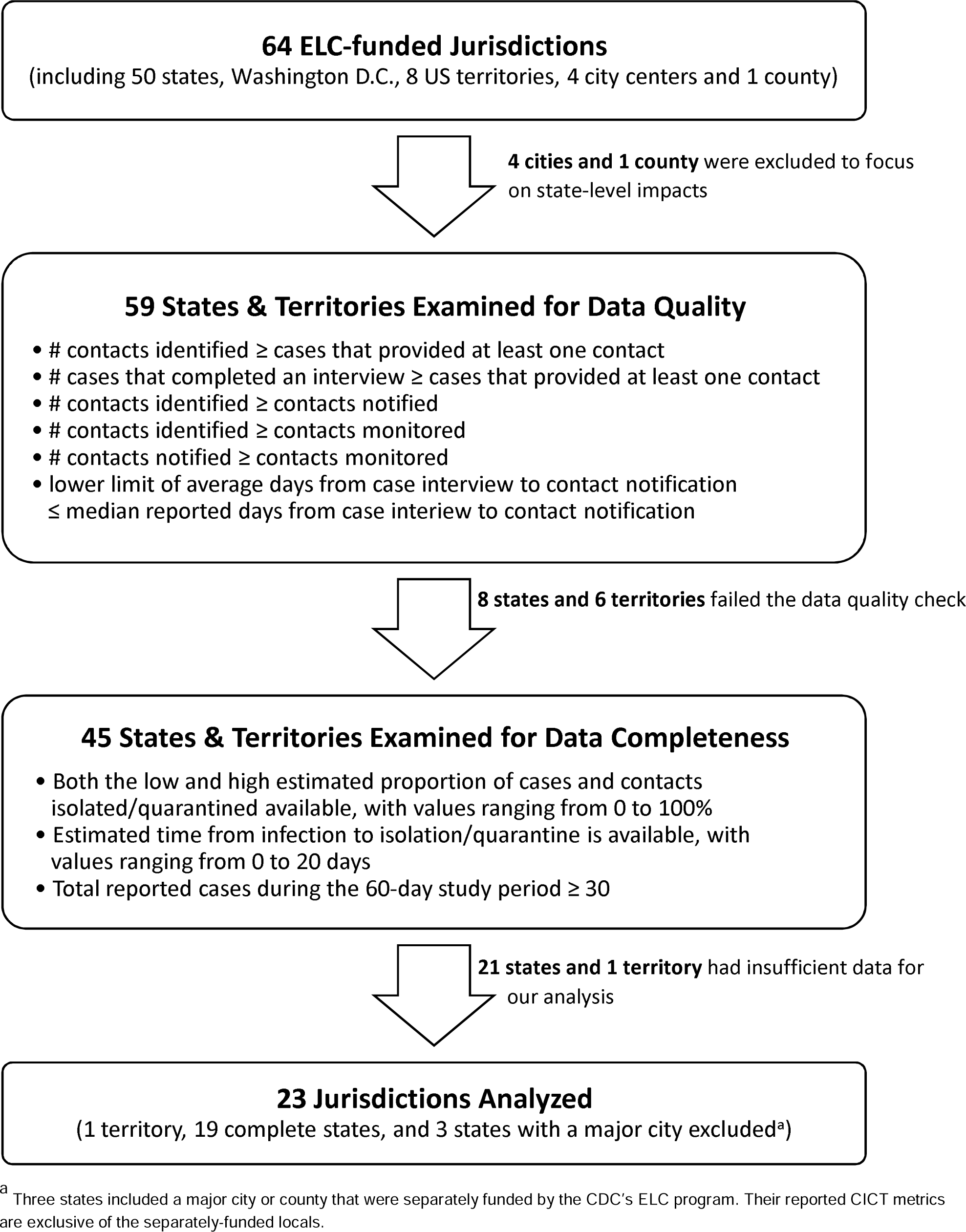
Inclusion and Exclusion Criteria for Analysis of Jurisdictions.

### Additional Results and Commentary

**eFigure 4.**
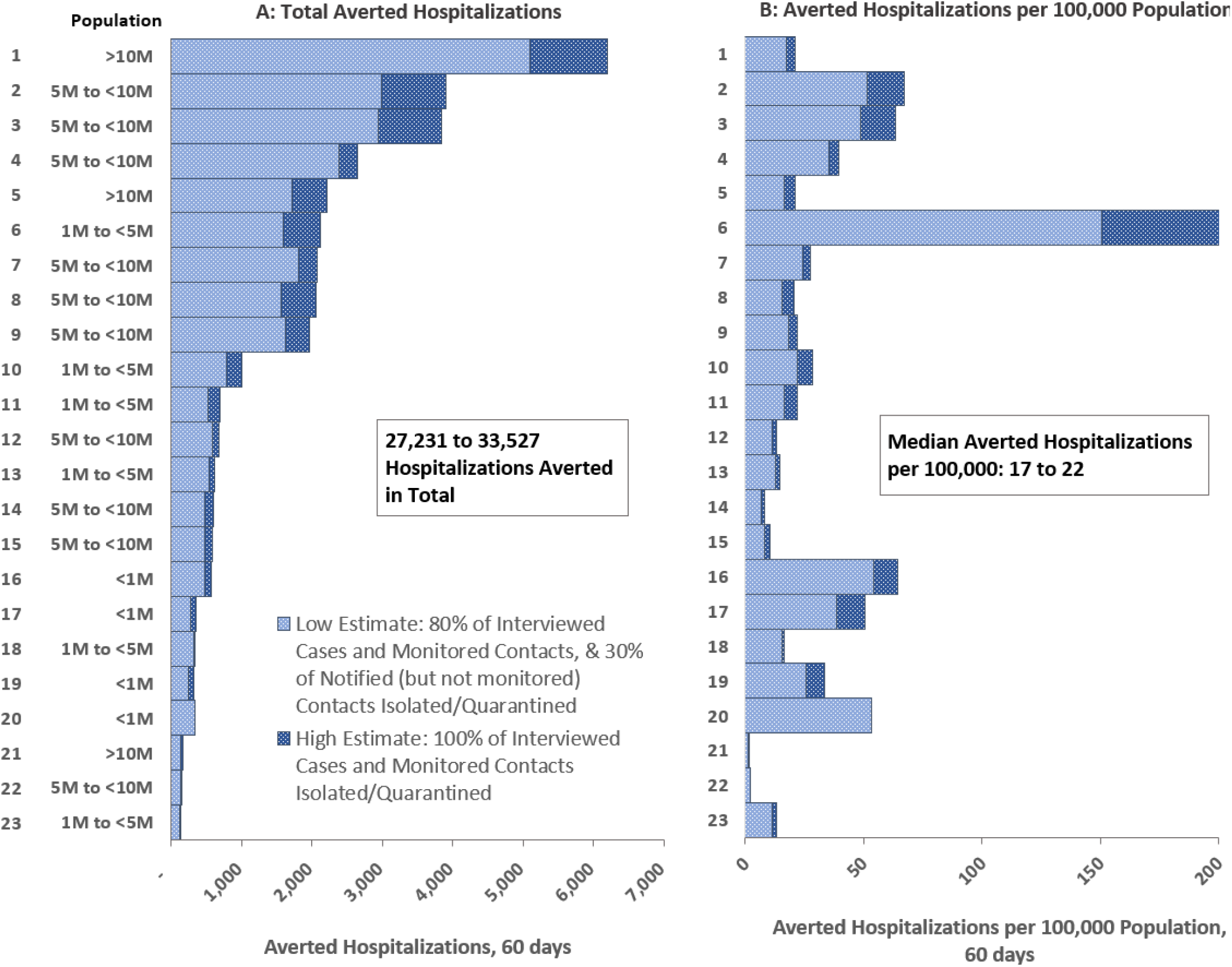
Estimated hospitalizations averted due to CICT programs from 11/25/20 – 1/23/21 (60 days)

**eTable 4.**
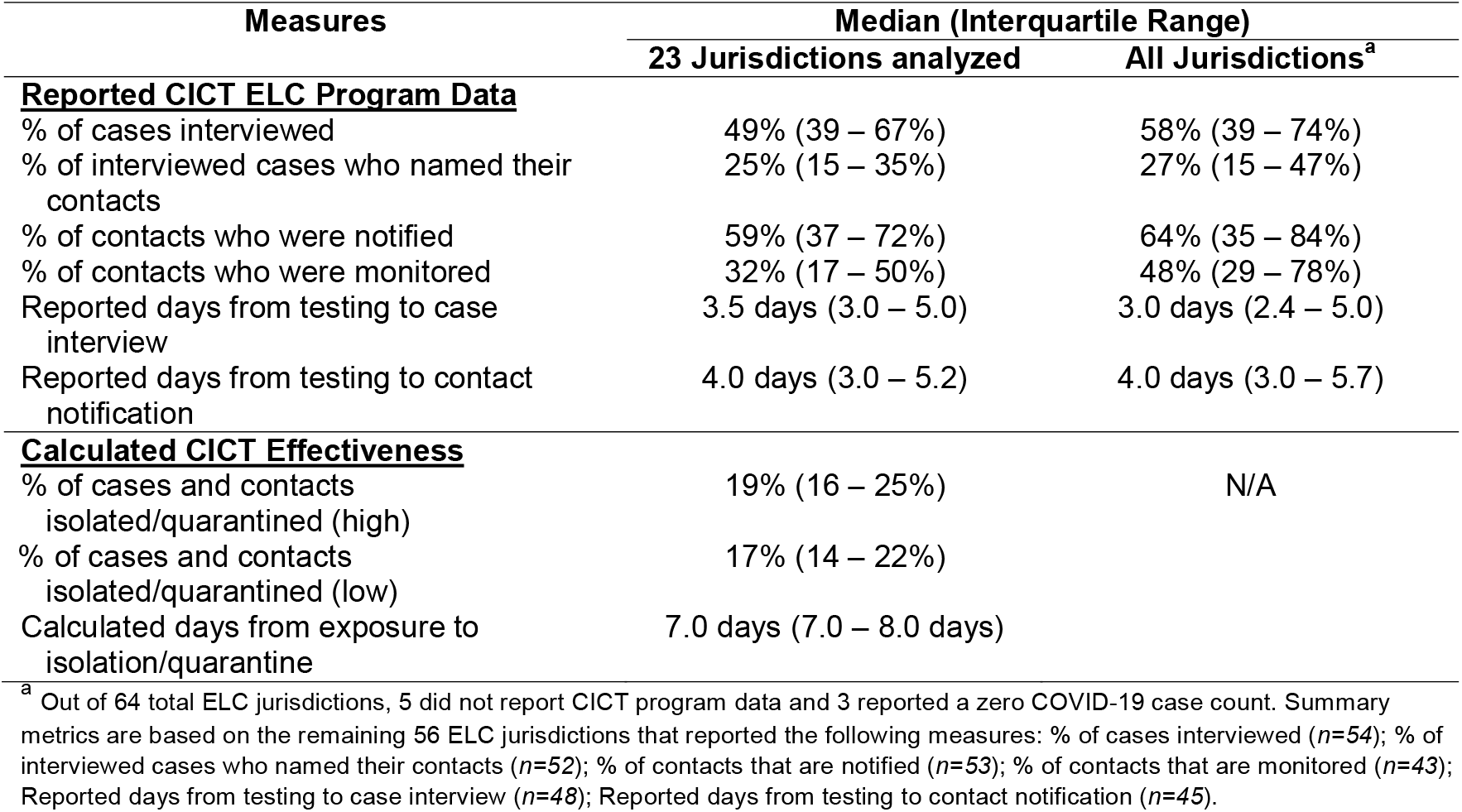
Summary of reported case investigation and contact tracing (CICT) data reported to CDC’s ELC program and calculated CICT effectiveness for the 23 jurisdictions analyzed and all funded jurisdictions, 11/25/20–12/24/20 (30 days)

**eTable 5.**
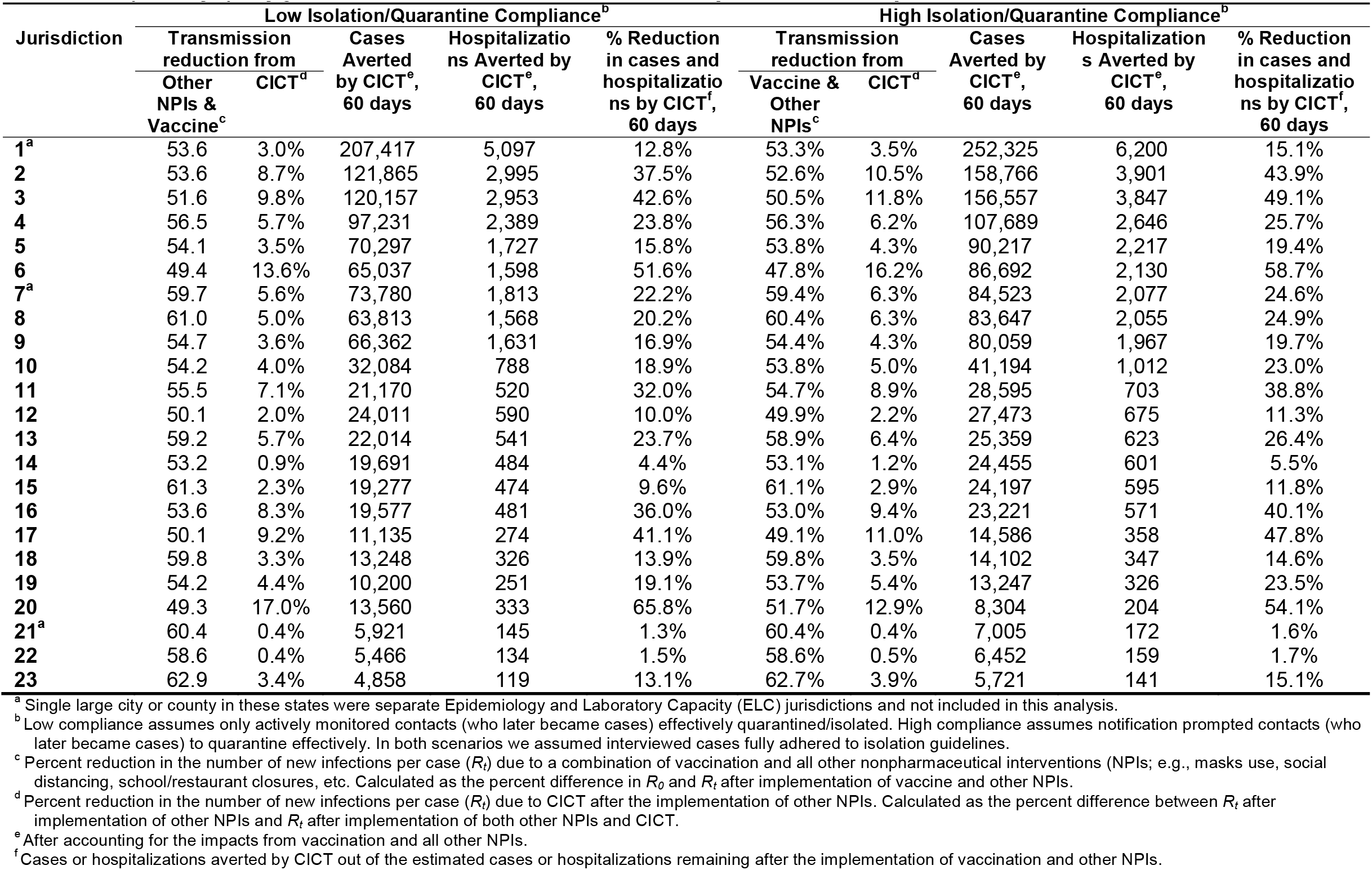
Estimated impacts of case investigation and contact tracing (CICT) and other interventions from 11/25/20 – 1/23/21 (60 days), by jurisdiction and assumed isolation/quarantine compliance.

### Isolation/Quarantine Compliance

A review of multiple cross-sectional population surveys in the UK suggests 40-45% of people who had COVID-like symptoms self-reported fully complying with isolation guidance during their infectious periods.^11^ Another survey in the US found that 85% of respondents who had COVID-like symptoms or tested positive stayed home (according to CDC guidelines) except to get medical care.^12^ And a third survey, also in the US, found that 93% of adults said they would definitely (73%) or probably (20%) quarantine themselves for at least 14 days if told to do so by a public health official because they had the coronavirus (*i*.*e*., they were confirmed cases; not just exposed contacts).^13^

### Instructions for using COVIDTracer Advanced Special Edition to estimate the number of COVID-19 cases and hospitalizations averted by CICT

These instructions will guide a user how to use the Special Edition version of COVIDTracer Advanced tool to repeat the analysis described in this manuscript to estimate COVID-19 cases averted by case investigation and contact tracing activities. The Special Edition version of COVIDTracer Advanced is a modification of the publicly available tool on CDC’s website that enables users to assess the impact of CICT before vaccine was widely available. Additional modifications would be required if you intend to explicitly account for vaccinated individuals (*e*.*g*., decreasing susceptible population over time, decreased risk of hospitalization among vaccinated individuals, etc).

Readers seeking basic information about the model, data elements, and definitions should refer the COVIDTracer Advanced User Manual at: https://www.cdc.gov/coronavirus/2019-ncov/php/contact-tracing/COVIDTracerTools.html

COVIDTracer Advanced uses the Windows operating system (Microsoft Windows 2010 or higher) and Excel (Microsoft Office 2013 or higher).

**Before starting, complete the following**

1. Determine your 60-day study period. The first day of your study period is your “model start date.” This “model start date” will be referenced later in these instructions. For example, if you are interested in estimating cases and hospitalizations averted by CICT during the 60-day period from January 1-March 1, 2021, your “model start date” is January 1, 2021.
2. Obtain these data for the jurisdiction of interest:
  a. Total population
  b. Total cases as of the day before the model start date (In the example study period above, this is the total cases reported as of December 31, 2020.)
  c. Cases reported during the past 14 days (In the example study period above, this is the sum of cases reported from December 18 to 31, 2020.)
  d. The case trend during the past 14 days (*e*.*g*., increasing, plateaued, decreasing)
  e. Daily (*i*.*e*., incident) case counts for the 60-day study period
  f. The following case investigation and contact tracing program metrics. These metrics are meant to be representative of the 60-day study period. If you don’t have such data for the entire study period, you may base these metrics on a shorter period (*e*.*g*., 30 days or 4-weeks) from the model start date (and assume they are representative of the full 60 days):
    i. Number of days from exposure to case isolation and contact quarantine
    ii. Percent (%) of all cases successfully isolated and contacts quarantined
3. Open the COVIDTracer Advanced_SpecialEdition tool (Supplement 2)
  a. When opening the spreadsheet file, click the “Enable Macros” button for full functionality of the tool.
  b. Enable Excel “Solver Add-In.” **Instructions**: in Excel, click on File → Options → Add-ins → select “Analysis ToolPak” → click “Go” (not the “Ok” button) → select checkbox for “Solver Add-In” and click “Ok.”
  The Solver button, 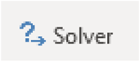 will appear in the “Data” menu.

**In worksheet, “A. Outbreak Details”**

**Step 1**: Enter the population for the jurisdiction of interest.

Enter the population of your jurisdiction
|  |  |  |
| --- | --- | --- |
| Total Population | 1,000,000 | persons |

**Step 2**: Enter the model start date, the total number of COVID-19 cases in the jurisdiction until the day before the model start date, and the number of cases reported in the last 14 days within the jurisdiction.

Enter information about case counts in your jurisdiction
|  |  |  |
| --- | --- | --- |
| Start Date | 1/1/2021 |  |
| Total Cases as of 12/31/2020 | 35,000 | cases |
| Cases in the last 14 days (from 12/18/2020 to 12/31/2020) | 5,000 | cases |

*Note: These data inputs will only create curves for the purpose of calculating resources needs. They are not intended as, nor should be interpreted as, forecasts of future cases*

**Step 3**: Set the pattern of daily cases over the past 14-day period selected in Step 3.

The default is “Daily case counts are slowly increasing.” However, if daily case counts have been changing rapidly, remaining constant, or decreasing over the last 14 days, select from the pull-down menu the pattern that best matches the jurisdiction’s data.

Enter estimates about the effectiveness of community interventions implemented through the present date
|  |  |
| --- | --- |
| Are community interventions currently in place? | Yes |
| Pattern of change in daily case counts over the last 14 days | Daily case counts are slowly increasing |

How to choose?

**In worksheet, “Case Counts”**

**Step 4**: Paste the jurisdiction’s daily case counts (*i*.*e*., incident cases) for the 60-day study period into the “Daily” column (column AH)

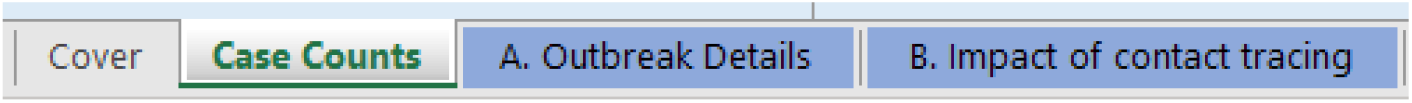

**In Worksheet, “B. Impact of Contact Tracing”**

**Step 5**: Using your representative CICT program data, enter values for:

- Number of days after infection that case is isolated
- % of all cases successfully isolated and contacts traced and monitored

| Contact Tracing Strategy Input | No Contact Tracing | Continued Contact Tracing | Strategy 3 (Optional) |
| --- | --- | --- | --- |
| Number of days after infection that case is isolated | 15 | 9 |  |
| % of all cases successfully isolated and contacts traced and monitored | 0% | 20.0% |  |
| Strategy Trigger | Symptoms | Contact ID |  |
| Contacts are identified and listed? | No | Yes |  |
| Contacts follow-up occurs? | No | Yes |  |

*\*Successfully traced = the strategy worked as assumed and transmission to the next generation was prevented*

**Step 6:** Estimate the % reduction in transmission due to community interventions (shown in cell G28) by fitting COVIDTracer Advanced’s simulated curve to your observed case curve. You will use the Solver Add-in to do this: The Solver Add-in finds an optimal solution for the % reduction in transmission due to community intervention by minimizing the mean squared error (a mathematical value describing the differences between both curves; shown in cell O32).

### Instructions for using the Solver

From the Excel menu tab, click “Data” and the “Solver” button, then follow the instructions described here to set up the parameters in the pop-up dialogue box (see screen shot below):

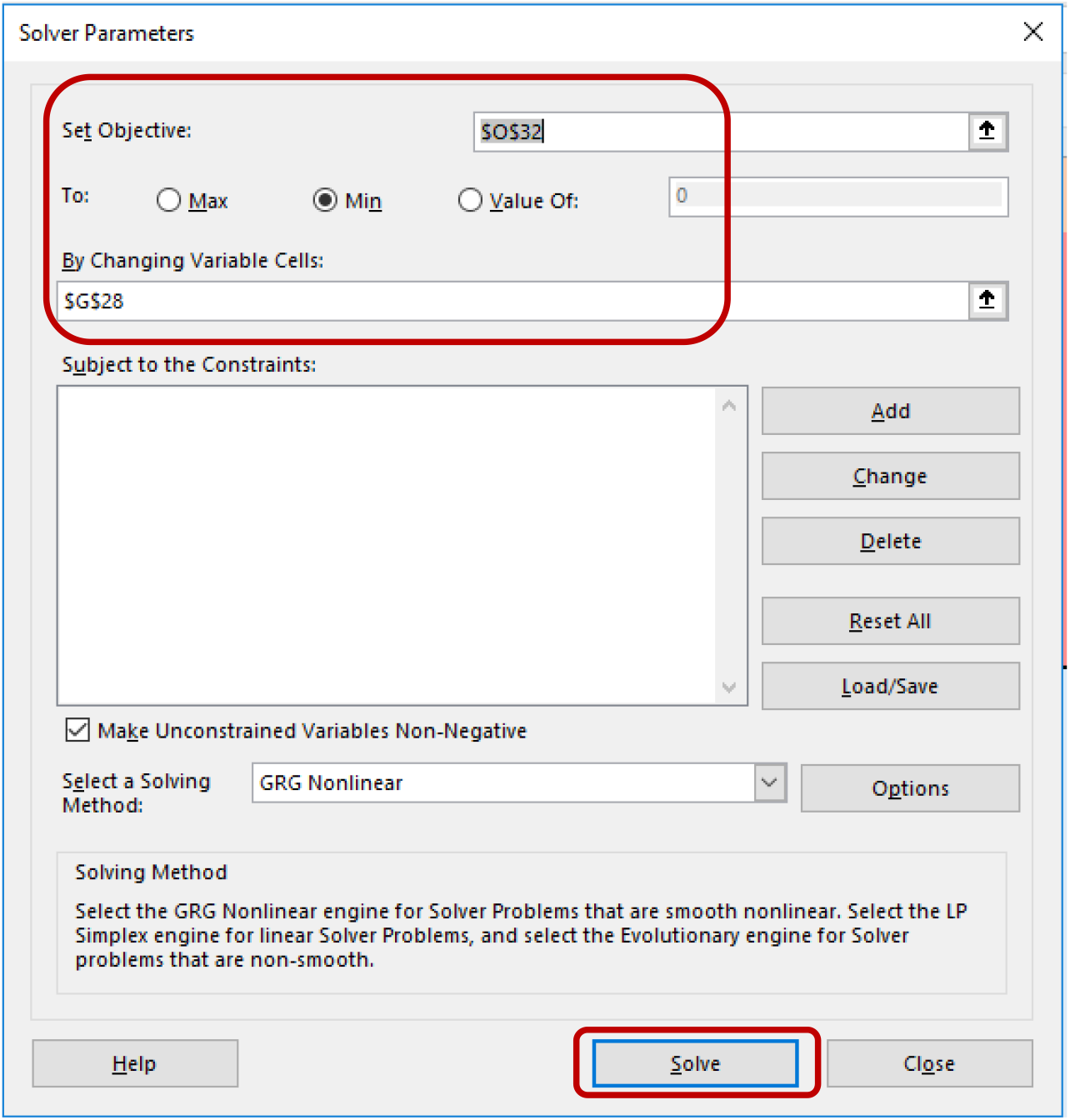

**Set Objective**: Set objective to cell “$O$32”, which is the mean squared error.

**To**: Select “Min”.

**By Changing Variable Cells**: Enter $G$28 (This cell refers the Solver to the “Estimated % reduction in transmission due to continued community interventions.”)

**Select a Solving Method**: For simplicity, we recommend selecting “GRG Nonlinear” from the drop-down menu.

**Click “Solve” button.**

Then the Excel Solver function will automatically find the optimal value (estimated % reduction in transmission due to continued community intervention) and populate the value in cell G28. The figure below shows a fitted curve (solid line) generated by COVIDTracer Advanced after Step 6, that minimizes deviation from the reported case counts (dashed line).

**Example Figure: Fitted curve using COVIDTracer Advanced**

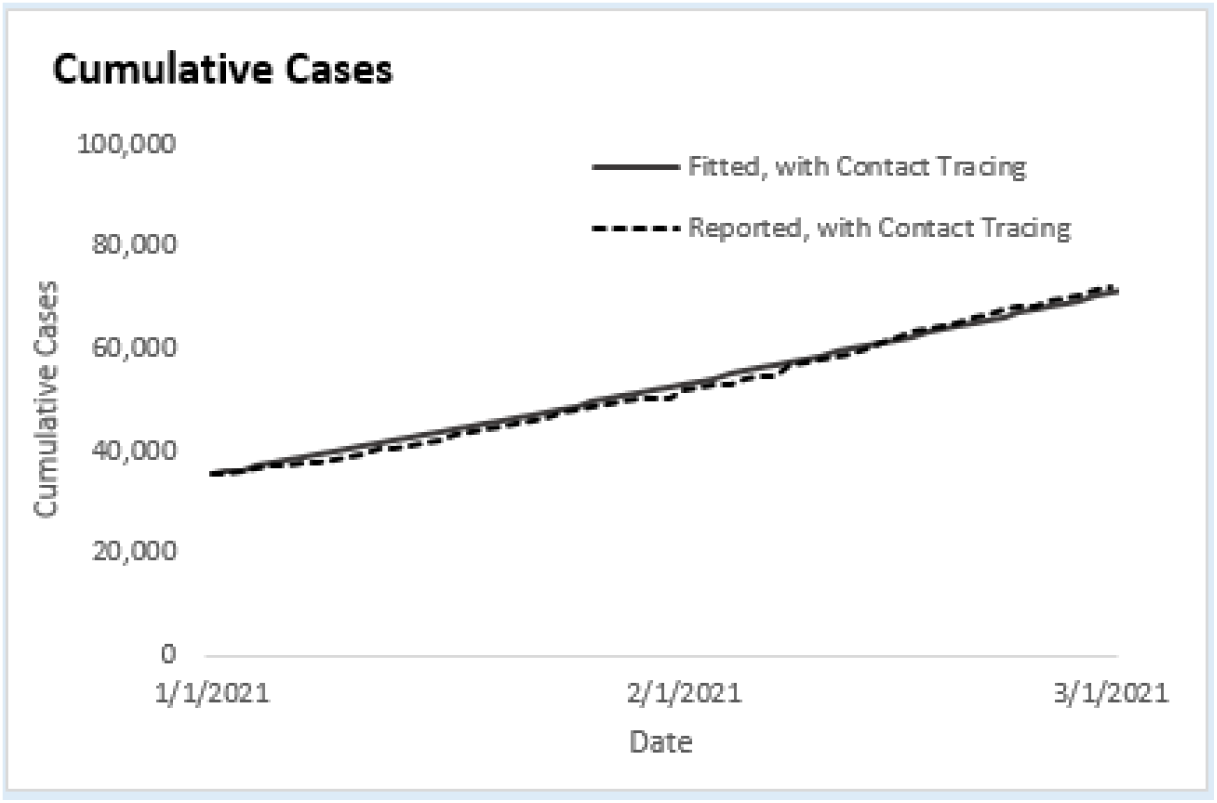

**In Worksheet, “Results – Cases Averted”**

**Step 7**. Users can find the % reduction in transmission due to CICT, and those that are attributable to all other interventions. The estimated number of cases and hospitalizations averted by CICT are also provided on this page.

Transmission Fraction
|  |  |
| --- | --- |
| **Transmission Reduction from Contact Tracing** | 4.5% |
| **Transmission Reduction from All Other Interventions** | 54.7% |
| **Remaining Transmission\*** | 43.3% |

\* Calculated as follows: $(1 - \text{reduction from CT}) * (1 - \text{reduction from other interventions})$

|  |  |
| --- | --- |
| **Cases Averted by Contact Tracing** | 8,937 |
| **Cases Averted per 100,000 population** | 894 |
| **% of Additional Cases Averted by Contact Tracing\*\*** | 19.7% |

\*\* Additional cases averted by contact tracing out of every 100 remaining cases after accounting for the impact of all other interventions (e.g., vaccination, facemask policies, social distancing).

|  |  |
| --- | --- |
| **Hospitalizations Averted by Contact Tracing** | 220 |
| **Hospitalizations averted per 100,000 population** | 22 |
| **% of Additional Hospitalizations Averted by Contact Tracing\*\*\*** | 19.7% |

\*\*\* Additional hospitalizations averted by contact tracing out of every 100 remaining hospitalizations after accounting for the impact of all other interventions.

## Footnotes

1 See *e*.*g*., 45 C.F.R. part 46, 21 C.F.R. part 56; 42 U.S.C. §241(d); 5 U.S.C. §552a; 44 U.S.C. §3501 et seq.

2 Isolated 100% of the time over the remaining duration of the recommended isolation/quarantine period.

3 Some cases can be identified before being symptomatic (e.g., during screening for various reasons)

## Notes

**Conflicts of Interest:** No conflicts to report

### Competing Interest Statement

The authors have declared no competing interest.

### Funding Statement

This work was performed as part of the official duties of all participants in support of the US CDC's COVID-19 Response.

